# Daughter circumcision and maternal life satisfaction: A cultural moderation effect revealed across two studies

**DOI:** 10.1101/2023.10.11.23296885

**Authors:** Paul S. Strand, Providence Maxwell, Justin Trang

**Affiliations:** Washington State University

## Abstract

**Introduction:** The present paper explored the relationship between maternal life satisfaction and the intergenerational transmission of female genital cutting (FGC; female circumcision).

**Methods:** Across two studies with more than 85,000 participants in 15 countries, maternal surveys reveal that the association is positive and moderated by country-level FGC prevalence.

**Results:** Contrary to predictions, in countries in which FGC is uncommon, it is more positively associated with maternal life satisfaction; and in countries in which it is common, it is weakly or negatively associated with maternal life satisfaction.

**Conclusion:** Results suggest a diversity of social motives for FGM. Customized messaging to reduce its inter-generational transmission should be considered.

**KEY MESSAGES:** *What is already known about this topic?:* Female genital cutting (FGC) has negative implications for health and gender equality and is practiced to different degrees in countries in parts of Africa, Asia, the Middle East, and their diasporas.

*What this study adds?:* This study explored the intergenerational transmission of FGC—in particular, how daughter FGC relates to maternal life satisfaction. Contrary to expectations, life satisfaction ratings were higher for mothers of circumcised daughters, although this relationship was moderated by country-level FGC frequency. In countries in which it is more common, daughter FGC is less strongly or negatively associated with maternal life satisfaction.

*How might this study affect research, practice, or policy?:* Study findings may inform anti-FGC messaging. In countries in which FGC is common, messaging should highlight its association with relative maternal dissatisfaction. In countries in which it is uncommon, messaging should highlight alternatives to FGC as an expression of cultural identity.

## INTRODUCTION

Female genital cutting (FGC), also known as female genital mutilation or female circumcision, involves the laceration of genital anatomical structures or the stitching of the vaginal orifice. The practice exists in parts of Africa, the Middle East, and Southeast Asia, usually undertaken in the context of religious or coming-of-age rituals with the aim of signaling or facilitating chastity, purity, and emasculated femininity (Andro et al., 2016; Hernlund & Shell-Duncan, 2020). It is most common in the rural and traditional areas of countries within which it is practiced (Population Reference Bureau, 2020).

In 2012, the United Nations General Assembly adopted a resolution to ban this practice (Johnson, 2012). The rationale being: (1) it is unsound from a reproductive health perspective and (2) it is a form of discrimination against women and girls contributing to gender inequality and constricted social and economic opportunities for them (Andro et al., 2016). Despite efforts to eradicate it the practice persists to the present day, albeit with somewhat decreasing frequency over time (Farouki et al., 2022).^1^

Social and religious norms are thought to be the basis of FGC (Ahinkorah et al., 2020; Andro et al., 2016; Berg & Denison, 2013; Hernlund & Shell-Duncan, 2020). Consistent with this, analysis of data collected by United Nations International Child’s Emergency Fund (UNICEF) across countries in which FGC is practiced revealed that the likelihood of a girl being cut is higher if her mother was cut. In addition, the likelihood of a woman or girl being cut increases with the proportion of cut women in her community (Population Reference Bureau, 2020). These findings suggest that the practice reflects community-level normative behavior that is intergenerationally transmitted—perhaps because the practice is socially reinforced and therefore rewarding within the socio-cultural contexts in which it is common.

While FGC has been investigated with respect to the influence of family practices and cultural norms, psychological factors have less often been investigated. If the practice is rewarding to those who condone or engage in it, one might expect that it would be associated with life satisfaction or happiness. That is, perhaps the motivation for individual families to engage in the practice with their daughters is a psychological one—it is consistent with increased happiness or life satisfaction of significant others. If maternal happiness derives from its alignment or compliance with social or cultural norms, then FGC should be most strongly associated with life satisfaction in countries in which the practice is more common.

This possibility is evaluated in the present paper. We explore whether maternal life satisfaction (MLS) is associated with the intergenerational transmission of FGC (as reflected in daughter FGC); and whether that relationship is moderated by country-level FGC. This constitutes an exploration of the notion that “FGM/C is sustained across generations in part by expectations and norms within a social or religious group at the community level. People living in proximity are more likely to influence each other’s expectations of appropriate behaviour” (Population Reference Bureau, 2020). We further explore if those associations reflect the impact of normative behavior on MLS. Support for this hypothesis would take the form of stronger daughter FGC—MLS associations in countries characterized by higher country-level FGC. If not, then the impact of country-level norms on the intergenerational transmission of FGC would seem to be independent of MLS.

Testing this hypothesis requires controlling for the influence of several possible confounds. Such confounds include individual-level and country-level variables that may themselves be associated with MLS or daughter FGC. These include mother education level, household wealth, and country-level wealth. For instance, the proposed moderating influence of country-level FGC may simply reflect differences in country-level wealth. Including country-level wealth as a variable within relevant statistical models would allow for examining the effect of country-level FGC on the association between daughter FGC and MLS that is unique from that of country-level wealth. At the within-family level it is necessary to similarly control for mother education and household wealth, both of which are likely associated with a cultural practice such as daughter FGC and with MLS. Finally, mother FGC is also included as a covariate predictor, thereby allowing for an estimate of the association between MLS and the intergenerational transmission of FGC, controlling for mother’s own personal experience of FGC on MLS.

## METHODS

### Procedure

The present study utilized data collected by UNICEF as part of its Multiple Indicator Cluster Surveys (MICS) project (https://mics.unicef.org/). MICS is the largest source of internationally comparable data on children and women worldwide with data collected in 120 countries, across 28 years, with 7 distinct rounds of data collection (UNICEF, nd a). We utilized data from MICS 5 and MICS 6 collected, respectively, between 2013-2016 and 2017-2022. Data for the present study included responses from adult women who self-identified as mothers, in countries for which daughter FGC data were available. Data were collected by trained interviewers in a private or semi-private one-to-one interview setting.

MICS data are de-identified, publicly available, and organized by country (https://mics.unicef.org/). Data for the present study existed in two separate files: a file labeled “FG” contained daughter circumcision data and all other study variables were contained in the “WM” file. Data preparation involved merging variables across these two files for each country and then merging cases across countries (UNICEF, nd b). In this way, final datasets were generated for MICS 5 and MICS 6, analyzed as Study 1 and Study 2, respectively.

### Participants

Participants for the present study were female offspring of women interviewed as part of the MICS data collection effort for whom valid responses existed for daughter FGC. Participation was further limited to those offspring for which valid responses existed for the dependent variable of the present study, maternal life satisfaction (MLS). Table 1 presents frequencies for these two variables for each country and the total sample. Also presented is the number of cases for which valid responses existed for both variables; those cases comprise the final sample for the present study. Additional columns in Table 1 provide information on the percentage of participants who experienced FGC and country-level per capita GDP. Additional demographic characteristics were as follows: average age of mothers, 30-34 years; average number of daughters per mother, Study 1: 2.61 (1.37), Study 2: 2.55 (2.51). daughter average age at circumcision, Study 1, 1.58 years (2.45), Study 2, 2.14 years (3.07).

**Table 1.** Per Country Participant Information.

| Table 1. Per Country Participant Information. |  |  |  |  |  |
| --- | --- | --- | --- | --- | --- |
| Participating Country | FG cases* | MLS cases<sup>^</sup> | Final Sample<sup>#</sup> | Daughter FGC Frequency<sup>+</sup> | GDP per capita |
| **Study 1: MICS 5** |  |  |  |  |  |
| Benin | 9722 | 1073 | 1059 | 0.3% | \$4056 |
| Cote d' Ivorie | 19840 | 2676 | 2676 | 12.9% | \$6538 |
| Guinea | 8938 | 10397 | 1257 | 45.4% | \$3187 |
| Guinea-Bassau | 8742 | 1342 | 1342 | 28.8% | \$2190 |
| Kenya | 2245 | 288 | 288 | <0.1% | \$5704 |
| Mali | 15205 | 2297 | 2253 | 77.4% | \$2517 |
| Mauritania | 12855 | 1458 | 1457 | 51.2% | \$6424 |
| Nigeria | 17,078 | 207 | 207 | 23.9% | \$5860 |
| Senegal | 5660 | 384 | 383 | 2.5% | \$9209 |
| Total (Average) | 100,285 | 10,987 | 10,922 | (11.5%) | (\$5,076) |
| **Study 2: MICS 6** |  |  |  |  |  |
| C. African Rep. | 15274 | 19074 | 15234 | 1.8% | \$967 |
| Gambia | 12417 | 16357 | 12411 | 36.7% | \$2510 |
| Ghana | 7357 | 10840 | 7357 | 0.4% | \$6498 |
| Guinea-Bissau | 8536 | 11142 | 8534 | 19.9% | \$2190 |
| Iraq | 9090 | 3046 | 2086 | 0.6% | \$10,862 |
| Nigeria | 17772 | 24825 | 17767 | 8.8% | \$5860 |
| Sierra Leon | 13541 | 18197 | 13532 | 6.4% | \$1931 |
| Togo | 4510 | 6045 | 4509 | 0.3% | \$2608 |
| Total (Average) | 88,497 | 109,526 | 81,430 | (9.4%) | (\$4178) |
| *Notes.* *FG Cases = cases for which a valid response existed to the “daughter circumcised” question. ^MLS Cases = cases for which a valid response existed for the MLS question. #Final Sample = cases for which a valid response existed for both questions. +Percentage of Final Sample cases for which response to “daughter circumcised?” was, yes. |  |  |  |  |  |

The study was reviewed and deemed exempt by the Washington State University Institutional Review Board (IRB#20245-001). The study was not conducted or conceived of with public or patient input.

### Measures

Maternal life satisfaction (happiness) was assessed with a single item: “Taking all things together, would you say you are very happy, somewhat happy, neither happy nor unhappy, somewhat unhappy, or very unhappy?” A “smiley card” served as a visual prompt depicting five facial icons ranging from upturned, smiling mouth (*very happy* = 1) to a downturned mouth (*very unhappy* = 5). Participants could verbalize or point to the relevant response. For the present paper, responses were reverse coded so that higher scores reflected happiness.

Daughter FGC was assessed by a single dichotomous question asked of mothers for each of their daughters: “Has she [daughter] been circumcised?” Mother FGC was assessed with a single dichotomous question addressed to mothers: “Have you been circumcised?” Valid response options were *yes* = 1, *no* = 2. For the present study, responses were reverse coded so that higher scores reflected, circumcised.

Mother education was assessed with a single item: “What is the highest level and grade or year of school you have attended?” Response options included: *without formal education* = 0; *primary education* (grade one to five) = 1; *secondary education* (grade six to ten) = 2; *higher secondary education and above* (grade 11, 12 and above) = 3.

Household Wealth scores were from the MICS Wealth Quintile Index (Martel, 2016). Quintiles were assigned to households in the entire MICS dataset for each country. They were generated using principle components analysis based on the following factors: ownership of household goods and amenities; persons per sleeping room; type of floor, roof, wall, cooking fuel, and sanitary facility; and source of drinking water.

Country-level Wealth was estimated for each country using *Gross Domestic Product at purchasing power parity per capita* (GDP), which improves upon nominal GDP by accounting for relative cost of living (https://www.worldometers.info/gdp/gdp-per-capita/). GDP values for 2022 were used and are reported for each country in Table 1.

Country-level Daughter FGC frequency was calculated for each country as the percentage of daughters in the final sample for each country for whom their mother identified as having been circumcised. Values for each country are reported in Table 1.

## RESULTS

### Data Analysis Plan

We utilized moderation analyses to investigate if the association between MLS and daughter FGC differed as a function of country-level daughter FGC rates. The statistical model generated to evaluate the primary hypothesis of the present study involved utilizing MLS as the dependent variable, daughter FGC as the independent variable, country-level FGC as the moderator variable, and the following variables as covariates: maternal education, household wealth, maternal FGC, and country-level wealth. Analyses were conducted using the PROCESS macro for Statistical Package for the Social Sciences (SPSS; Hayes, 2022).

The primary prediction of the present study is that daughter FGC will be a more positive (or less negative) predictor of MLS in countries with higher within-country FGC frequencies. This would confirm that conforming to cultural norms with respect to FGC is associated with life satisfaction for mothers, which would provide a psychological-level explanation for the intergenerational transmission of FGC within countries within which it is prevalent.

Results are organized into two studies. Study 1 reporting on results for the MICS 5 dataset and Study 2 the MICS 6 dataset. Study 2 represents a direct replication of Study 1. Missing data were treated as missing at random and listwise deletion procedures were utilized.

### Study 1 Results

Table 2 provides means and standard deviations for all individual-level variables along with their bivariate correlations. MLS was positively associated with education and, surprisingly, negatively associated with household wealth. MLS was also positively associated with mother and daughter circumcision. Household wealth and education were negatively associated with both mother and daughter circumcision. Mother circumcision was a significant predictor of daughter circumcision.

**Table 2.** Correlations and Means and Standard Deviations for individual-level variables for Study 1 and Study 2.

|  |  |  |  |  |  | Study 1 | Study 2 |
| --- | --- | --- | --- | --- | --- | --- | --- |
| Variable | 1 | 2 | 3 | 4 | 5 | M (SD) | M (SD) |
| 1. MLS | -- | .06** | -.04** | .09** | .05** | 4.31 (8.83) | 3.88 (1.09) |
| 2. Education | .03** | -- | .43** | -.21** | -.16** | 1.69 (0.91) | 0.92 (1.04) |
| 3. Wealth | .05** | .41** | -- | -.10** | -.08** | 2.91 (1.39) | 2.84 (1.39) |
| 4. M_FGC | .04** | -.24** | -.12** | -- | .54** | .58 (.49) | .45 (.50) |
| 5. D_FGC | .09** | -.16** | -.08** | .42** | -- | .31 (.46) | .14 (.35) |

*Notes.* Study 1 correlations are above the diagonal and Study 2 correlations are below the diagonal. MLS = Maternal life satisfaction; M\_FGC = Mother female genital cutting (dichotomous); D\_FGC = Daughter female genital cutting (dichotomous).
\* $p < .01$ , \*\* $p < .001$

Table 3 provides the results of the regression estimates for all variables entered in the moderation analysis for which MLS is the dependent variable. All entered variables were significant predictors except Household wealth. Model summary statistics were as follows, *R* = .208, *R*^2^ = .043, MSE = .664, *F* (7, 10,772) = 69.65, *p* < .001. For the interaction term (Daughter FGC x Country-level FGC) the test of the highest order unconditional interaction revealed Δ*R*^2^ = .008, *F* (1, 10,772) = 94.99, *p* <.001.

**Table 3.** Results of regression analyses for variables predicting MLS for Study 1 and Study 2.

|  |  |  |  |  |
| --- | --- | --- | --- | --- |
| Predictor | Coefficient | SE | *t* | 99% CI |
| **Study 1** |  |  |  |  |
| Maternal Education | .045 | .010 | 4.675** | .020 - .069 |
| Family Wealth | -.020 | .006 | -3.223 | -.037 - -.004 |
| Country GDP | .000 | .000 | 12.144** | .000 - .000 |
| Country FGC Frequency | .006 | .000 | 12.519** | .005 - .007 |
| M_FGC | .070 | .022 | 3.230** | .014 - .126 |
| D_FGC | .425 | .050 | 8.657** | .295 - .555 |
| D_FGC x Country FGM Frequency | -.008 | .001 | -9.746** | -.011 - -.006 |
| **Study 2** |  |  |  |  |
| Maternal Education | .017 | .004 | 10.808** | .006 - .027 |
| Family Wealth | .045 | .003 | 14.79** | .037 - .052 |
| Country GDP | .000 | .000 | 43.911** | .000 - .000 |
| Country FGC Frequency | .020 | .000 | 51.378** | .019 - .021 |
| M_FGC | .099 | .009 | 11.538** | .077 - .121 |
| D_FGC | .329 | .022 | 14.850** | .273 - .387 |
| D_FGC x Country FGM Frequency | -.015 | .001 | -17.338** | -.017 - -.012 |
| Notes. *$p < .01$; **$p < .001$ | | | | |

Table 4 provides the results of the moderation analysis evaluating conditional effects of country-level FGC frequency on the association between Daughter FGC and Maternal Life Satisfaction. As shown in Table 4 (Study 1 results), Daughter FGC is significantly related to MLS and country-level FGC frequency significantly moderated that relationship. This interaction is illustrated in Figure 1. The interaction was probed by testing the conditional effects of Daughter FGC on MLS at three levels of country-level FGC frequency, at the 16^th^ percentile, the 50^th^ percentile, and the 84^th^ percentile. As shown in Table 4, Daughter FGC was significantly positively related to MLS at Country-level FGC values of 12.9 and 28.8 (the 16^th^ and 50^th^ percentiles, respectively) and significantly negatively related to MLS at a value of 77.4 (the 84^th^ percentile). The Johnson-Neyman technique showed that the relationship between Daughter FGC and MLS was significant at values of country-level FGC less than 43.54 and more than 56.97.

**Figure 1.**
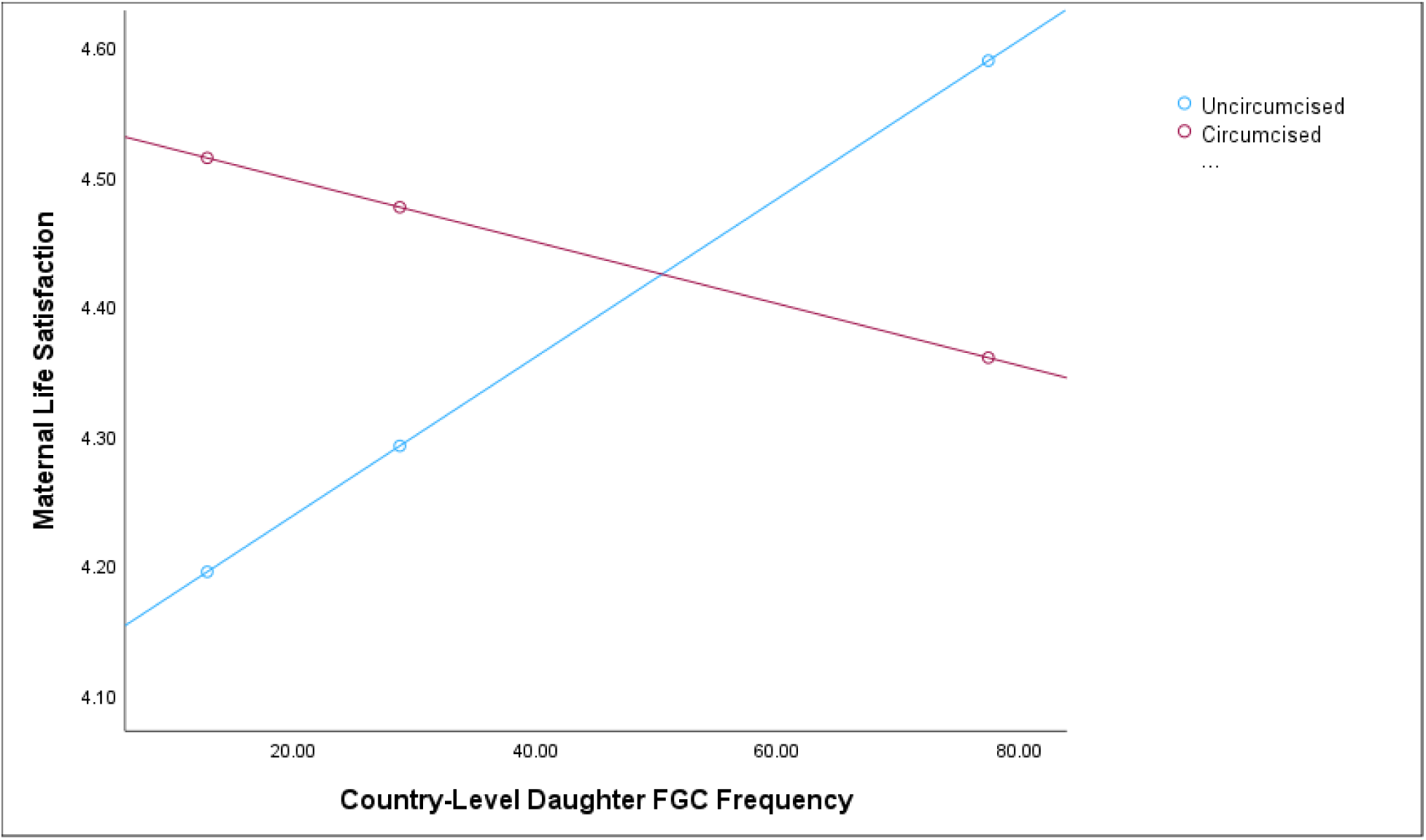
Maternal life satisfaction-daughter circumcision association by country-level daughter FGC frequency for Study 1.

**Table 4.** Conditional Effects of Daughter FGC on MLS at values of the moderator for Study 1 and Study 2.

| Moderator value | Coefficient | SE | *t* | 99% CI |
| --- | --- | --- | --- | --- |
| **Study 1** |  |  |  |  |
| 12.90 | .316 | .041 | 7.801** | .212 - .421 |
| 28.80 | .182 | .030 | 6.146** | .106 - .259 |
| 77.400 | -.227 | .030 | -7.618** | -.304 - -.150 |
| **Study 2** |  |  |  |  |
| 0.600 | .320 | .022 | 14.729** | .264 - .376 |
| 6.400 | .236 | .018 | 13.091** | .190 - .282 |
| 19.90 | .039 | .013 | 3.074* | .006 - .072 |
| Notes. *$p < .01$; **$p < .001$ | | | | |

### Study 1 Discussion

Study 1 results are contrary to predictions. The moderation analysis revealed a reversal effect whereby Daughter FGC and MLS were significantly associated with one another at all three levels of the moderator variable, country-level FGC. However, a reversal effect was found whereby in countries in which FGC is common Daughter FGC is associated with lower levels of MLS, and in countries in which FGC is uncommon Daughter FGC is associated with higher levels of FGC. This significant moderation effect was observed for a model controlling potential confounding variables including mother education, household wealth, and country-level wealth as measured by GDP. In addition, mother’s own history of FGC was also controlled for which ensures that the association between MLS and daughter FGC reflects the intergenerational relationship between those two variables controlling for mother’s own history of FGC.

These results suggest that the intergenerational transmission of FGC cannot be attributed to positive emotions stemming from conformity to cultural norms. Rather, the intergenerational transmission of the practice appears to occur despite its negative association with maternal happiness in countries in which it is common.

### Study 2 Results

Means and standard deviations for all individual-level variables, along with their bivariate correlations, are presented in Table 2. MLS was positively associated with education, household wealth, mother circumcision, and daughter circumcision. Household wealth and education were associated with MLS and daughter FGC. Mother FGC was a significant predictor of daughter FGC.

Table 3 provides the results of the regression estimates for all variables entered in the moderation analysis for which MLS is the dependent variable. All entered variables were statistically significant predictors. Model summary statistics were as follows, *R* = .252, *R*^2^ = .064, MSE = 1.11, *F* (7, 77,844) = 755.46, *p* < .001. For the interaction term (Daughter FGC x Country-level FGC) the test of the highest order unconditional interaction revealed Δ*R*^2^ = .004, *F* (1, 77,844) = 300.60, *p* <.001.

Table 4 provides the results of the moderation analysis evaluating conditional effects of country-level FGC frequency on the association between Daughter FGC and Maternal Life Satisfaction. As shown in Table 4 (Study 2 results), Daughter FGC is significantly related to MLS and country-level FGC frequency significantly moderated that relationship. This interaction is illustrated in Figure 2. The interaction was probed by testing the conditional effects of Daughter FGC on MLS at three levels of country-level FGC frequency, the 16^th^, 50^th^, and 84^th^ percentiles. As shown in Table 4, daughter FGC was significantly positively related to MLS at country-level FGC values of 0.60 (the 16^th^ percentile), 6.40 (50^th^ percentile), and 19.90 (the 84^th^ percentile), with a steady decrease in the differences at higher levels of country-level FGC. The Johnson-Neyman technique showed that the relationship between Daughter FGC and MLS was significant at values of country-level FGC less than 20.34 and greater than 24.90.

**Figure 2.**
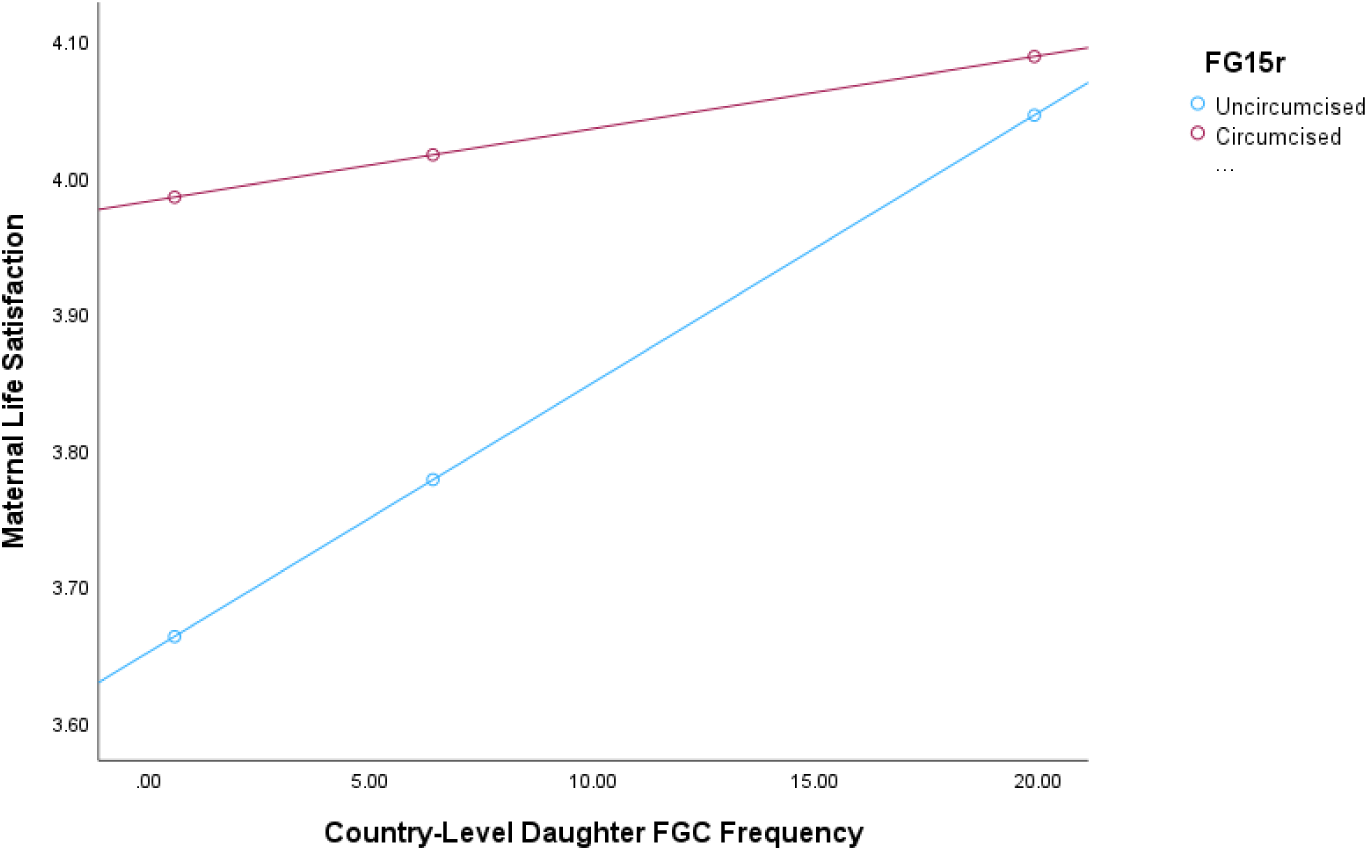
Maternal life satisfaction-daughter circumcision association by country-level daughter FGC frequency for Study 2.

### Study 2 Discussion

Consistent with the findings of Study 1, Study 2 results confirm that the association between Daughter FGC and MLS was lower, not higher, in countries characterized by higher levels of FGC. That is, daughter circumcision is more strongly associated with maternal happiness in countries in which the practice is uncommon. As such, the intergenerational transmission of the practice cannot be attributed to positive emotions stemming from cultural conformity.

## GENERAL DISCUSSION

The question arising from the results of these two studies is the following: Why, in countries in which daughter circumcision is culturally normative, are mothers of circumcised daughters less happy than mothers of uncircumcised daughters? Such findings are contrary to the notion that cultural conformity is personally rewarding and the impetus for its perpetuation.

One possibility concerns the potential negative impact on the psychological well-being of exposure to community-level FGC. For instance, one prominent African sociologist, theologian, and expert on female circumcision identified her own opposition to the practice as arising from witnessing the suffering and carnage arising from the practice in her native Kenyan community (Wangila, 2015). Moreover, recent literature suggests that FGC may be associated with adverse mental health outcomes, although this conclusion is tempered by equivocal findings across generally low-quality studies (Abdalla & Galea, 2022; Lien, 2022; O’Neill & Pallitto, 2021).

Indeed, for both samples reported on here, levels of maternal life satisfaction were *higher* for mothers who reported having experienced FGC themselves and who reported it for their daughters. Nevertheless, it may be that cultural forces (perhaps coercive) overpower individual-level maternal aversion to the practice in societies in which it is common.

On the other hand, it could be that maternal unhappiness is associated with higher levels of compliance with cultural customs. If so, this effect is in addition to any influence toward cultural conformity of the individual- and country-level variables controlled for in the present study, including maternal education, household wealth, country-level wealth, and her own personal circumcision history. Of course, given the data are cross-sectional and not longitudinal, the present studies cannot disentangle issues of direction of effect, nor can they rule out unmeasured third variable effects. This constitutes a primary weakness of the present study.

In countries in which it is uncommon, FGC might symbolize a linkage to a threatened identity or heritage. For instance, the term, *ngaitana*, which means “I will circumcise myself”, became a slogan in response to national and international sanctions against the practice in Kenya (Wangila, 2015). As such, the practice of daughter FGC in cultural contexts in which it is uncommon may reflect agency against social norms perceived as threatening to self and culture. In that context, FGC may reflect or bring about a sense of personal agency and life satisfaction. Indeed, in describing those who defend the practice, Wangila (2015) notes that “the agency of the individual is becoming accepted in Kenyan communities as a way of resisting social norms [that are contrary to FGC]”. In sum, depending on the socio-cultural context, the practice of FGC may reflect different and even contradictory personal and social motivational forces.

The implications of these findings for efforts to reduce the practice of FGC is that these different motivational forces must be recognized and addressed. For instance, in countries in which FGC is common, it should be communicated that the practice does not bring maternal satisfaction. In countries in which FGC is uncommon, efforts should be concerned with identifying and supporting alternative forms of cultural self-expression, perhaps particularly among those who are minoritized or feel excluded from expressing their own cultural values and heritage.

Ours is the first study we are aware of that has explored how a psychological variable, maternal life satisfaction, relates to the intergenerational transmission of FGC. Future research should further explore the role of psychological factors as they relate to the intergenerational transmission of FGC. It is worth considering how the practice relates to the psychological well-being of nonmaternal family members. Indeed, little or no work has explored differences in cross-parent or cross-generational attitudes related to FGC. Similarly, it would be worth exploring how seemingly contradictory motives may act to maintain a cultural practice across different cultural contexts. It would also be useful to explore how life satisfaction ratings relate to the different forms of FGC (genital nicking versus labia stitching, for example), which may be differentially associated with disability and trauma (Abdalla & Galea, 2022). Regarding the psychological significance of FGC, it would be useful to explore how it may be in the service of cultural conformity or cultural nonconformity, depending on the cultural context.

## ENDNOTE

1. There is concern that the UN resolution against FGC constitutes cultural imperialism (Earp, 2022).

## Data Availability

Data are available upon request made to the first author.

